# A Randomized, Double-Blind, Placebo Controlled Trial to Assess the Effectiveness and Safety of Melatonin and Three Formulations of Floraworks Proprietary TruCBN™ for Improving Sleep

**DOI:** 10.1101/2023.11.07.23298230

**Authors:** Antonija Kolobaric, Jessica Saleska, Susan J Hewlings, Corey Bryant, Christopher S. Colwell, Christopher R. D’Adamo, Jeff Chen, Emily K Pauli

## Abstract

Introduction: Prescription sleep aids often have side effects, making alternative solutions necessary. Melatonin is an efficacious sleep aid and is often the comparator for alternatives. The phytocannabinoid cannabinol (CBN), has a potential mechanism of action but minimal evidence to support its effectiveness. Methods: TruCBN™ is a hemp-derived cannabinol (CBN) sleep aid. The primary aim of this randomized, double-blind, placebo-controlled study was to assess the safety and effects of three formulations of TruCBN™ [25 mg (n=206), 50 mg (n=205), 100 mg (n=203)], on sleep quality measured by PROMIS Sleep Disturbance 8A scale, relative to placebo (n=204). As a secondary aim, the effectiveness and safety of these formulations relative to a 4 mg melatonin (n=202) were assessed. Exploratory measures were stress (PROMIS Stress 4A), anxiety (Anxiety 4A), pain (PROMIS™ PEG) and well-being (WHO 5). Results: All treatment groups (25, 50 and 100 mg TruCBN™) and the 4 mg melatonin group experienced significant improvement in sleep quality relative to placebo. There were no significant differences between any of the TruCBN™ groups and melatonin for improving sleep. Participants taking 100 mg TruCBN™ showed a larger decrease in stress compared to the placebo group. There were no significant differences in anxiety, pain, well-being or frequency of side effects between any TruCBN™ group and placebo. There was no significant difference in improvements in sleep quality between any of the treatment groups and 4mg melatonin. Discussion: Orally ingested TruCBN™, 25 mg, 50 mg and 100 mg, is a safe and effective alternative for the improvement of sleep.

Sterling Institutional Review Board (SIRB) approved the study [10147-EKPauli]. The study was registered on ClinicalTrials.gov [NCT05511818] on August 21, 2023.

## Introduction

Sleep deprivation can have a profound impact on overall well-being, negatively impacting brain function,^1^ cognitive performance,^2^ emotional well-being,^2,3^ and physical health.^4,.5^ Inadequate sleep is associated with poor overall mental health as well as increased perceived stress and anxiety.^.6^ Individuals who are sleep deprived are less productive and report a lower overall quality of life.^7^Despite these consequences, about one in three American adults do not get sufficient sleep each night.^8^ Robust clinical evidence supports the use of prescription drug interventions, such as benzodiazepine receptor agonist drugs, for treating sleep disorders.^9,10^ Nevertheless, concerns persist as to whether they are effective in the long term and over their numerous side effects, including the considerable risk of abuse and dependence.11 Safe and accessible alternative therapies must be evaluated to improve the well-being of those suffering from sleep difficulties. Melatonin is a popular alternative to prescription sleeping medications with a large amount of clinical data to support its efficacy. Because of its widespread use and clinical effectiveness, melatonin is often the sleep aid to which other non-prescription interventions are compared.^12-14^

Cannabis preparations have also gained increasing attention as potential alternative therapies for addressing sleep disturbances.^15^ Cannabinoids exert diverse effects on the human body through their interaction with the endocannabinoid system (ECS), which is distributed throughout the brain, central nervous system, and peripheral nervous system.^16^ The presence of the ECS in the hypothalamic-pituitary-adrenal axis (HPA) and sympathetic nervous system support its role in regulating stress, feelings of anxiety and pain.^17^ Additionally, the ECS has been proposed to regulate the circadian sleep/wake cycle, suggesting that cannabinoids play a role in modulating sleep and potentially aspects of health that sleep impacts such as stress and anxiety.^18^ The effect of the common phytocannabinoids, delta-9 tetrahydrocannabinol (THC) and cannabidiol (CBD), on sleep has been studied and is well supported by the role of the ECS on circadian regulation.^19^ Cannabinol (CBN), a rarer phytocannabinoid, has rapidly grown in popularity as a sleep aid, with many manufacturers claiming that it has sleep-inducing effects.^20^ Yet, despite anecdotal evidence and a plausible mechanism of action via the ECS, there is limited research on the compound’s effect on sleep. Preclinical and anecdotal evidence suggests that CBN could prolong sleep,^21^ though to our knowledge no published blinded randomized placebo-controlled trials have studied CBN’s effectiveness for disturbed sleep without combining it with other ingredients. Rigorous large-scale clinical trials are needed to assess the dose, efficacy, and safety of CBN for sleep.

The primary aim of the study was to assess the safety and effects of three formulations of softgels containing varying amounts of TruCBN™ (25-50-100mg) on sleep quality, relative to placebo control. As a secondary aim, we also sought to assess the comparative effectiveness and safety of these TruCBN™ products relative to a softgel containing 4 mg melatonin. Furthermore, because of the interconnection between sleep, stress, anxiety, pain and overall well-being, and the potential effect that TruCBN™ may have on these indices because of its effect of the ECS, we sought to include the effect on these outcomes as exploratory measures.

## Methods

This study, Radicle^TM^ Rest, was a randomized, double-blind, placebo-controlled parallel trial designed to assess the effects of 3 formulations of TruCBN™ and 1 formulation of melatonin softgels on sleep, anxiety, stress, pain, and overall health-related quality of life. The study was decentralized; participants did not attend any in-person visits and all data were collected via online surveys which participants accessed via participant specific hyperlinks sent to them at scheduled times through their preferred means of communication (email or SMS text). Participants were recruited online from across the United States through social media, Radicle Science’s electronic mailing list, and a third-party consumer network with nationwide representation. Recruitment emails containing links to the study screener were sent to those within the Radicle Science mailing list and consumer network, while social media advertisements led to a study landing page with a link to the study screener. Participants were eligible if they were 21LJyears old or older, resided in the United States, expressed a desire for better sleep, and ranked their desire for better sleep as a primary reason for taking a dietary supplement. Individuals were excluded if they were pregnant or breastfeeding, or taking medications that posed a health risk when used in conjunction with any of the study product ingredients. Eligible individuals were directed to a secure online portal to provide informed consent. Participants indicated their consent electronically by signing the informed consent form and were sent a digital copy of the electronic consent. Eligible individuals were advised to consult with their healthcare provider before participating if they had a diagnosed medical condition, were on any prescription medication or supplements, or had any upcoming medical procedures planned. Immediately following informed consent, participants completed an intake survey which collected basic demographic information, health behaviors, and experienced sleep quality. This research process has been successfully implemented for several other dietary supplement clinical trials.^22,23^

Recruits who consented to participate and completed intake were randomized to one of five study arms (see below for details on randomization): (1) Softgel A (containing 4 mg melatonin), (2) Softgel B (containing 25 mg TruCBN™), (3) Softgel C (containing 50 mg TruCBN™), (4) Softgel D (containing 100 mg TruCBN™) and (5) Placebo control. All study products were provided by the partnering manufacturer. After receiving all study products at our warehouse, and before shipping them to participants, we sent samples from each arm to an independent laboratory to ensure active ingredient presence, potency, and lack of contaminants. Participants were instructed to take one softgel 1-2 hours before bedtime. The study was double-blind; neither the participants nor those who collected and analyzed the data were aware of the product participants received until the conclusion of the study. Upon receiving the product, participants were asked to verify the study product alpha-numeric identification coder on their product to ensure that they received their assigned product. For 5LJweeks following the study initiation and baseline health outcome assessment (4LJweeks after taking the study product and 1 week after finishing the study product), participants were asked to complete online surveys, which they accessed via unique hyperlinks sent at scheduled times via email or text. During the baseline week, participants completed health outcome assessments of their sleep, feelings of anxiety, stress, pain, and overall well-being, using validated, patient-reported outcome measures (Table 1).

**Table 1.** Validated measures for key outcomes used in Radicle Rest study.

| Measure | Description | Scoring interpretation | How was this collected? |
| --- | --- | --- | --- |
| PROMIS Sleep Disturbance 8a | 8-item measure assessing sleep disturbance (sleep | Scoring from 8 to 40, with higher scores translating to | All participants received this measure within their weekly |
|  | quality) in the past 7 days | greater sleep disturbance | health surveys |
| PROMIS Anxiety 4a | 4-item measure assessing frequency of anxiety symptoms in the past 7 days | Scoring from 4 to 20, with higher scores translating to greater anxiety | Participants who endorsed anxiety symptoms received this measure in their weekly health surveys. |
| PROMIS Stress 4a | 4-item measure assessing frequency of stress symptoms in the past 7 days | Scoring from 4 to 20, with higher scores translating to greater stress | Participants who endorsed stress symptoms received this measure in their weekly health surveys. |
| PEG (Pain, Enjoyment, General Activity) scale | 3-item measure assessing pain intensity and interference in the past 7 days | Scoring from 0 to 10, with higher scores translating to greater pain | Participants who endorsed pain symptoms received this measure in their weekly health surveys. |

Throughout the study duration, participants electronically received a health survey asking them to report their study product usage and health outcome assessments for their sleep disturbance, feelings of anxiety, stress, pain, and overall well-being from the past week using the same validated health measures used at baseline (Figure 1). In every study survey, following receipt of their product, participants were also prompted to report any side effects and were encouraged to contact the research team directly if they experienced side effects at any point.

**Figure 1.**
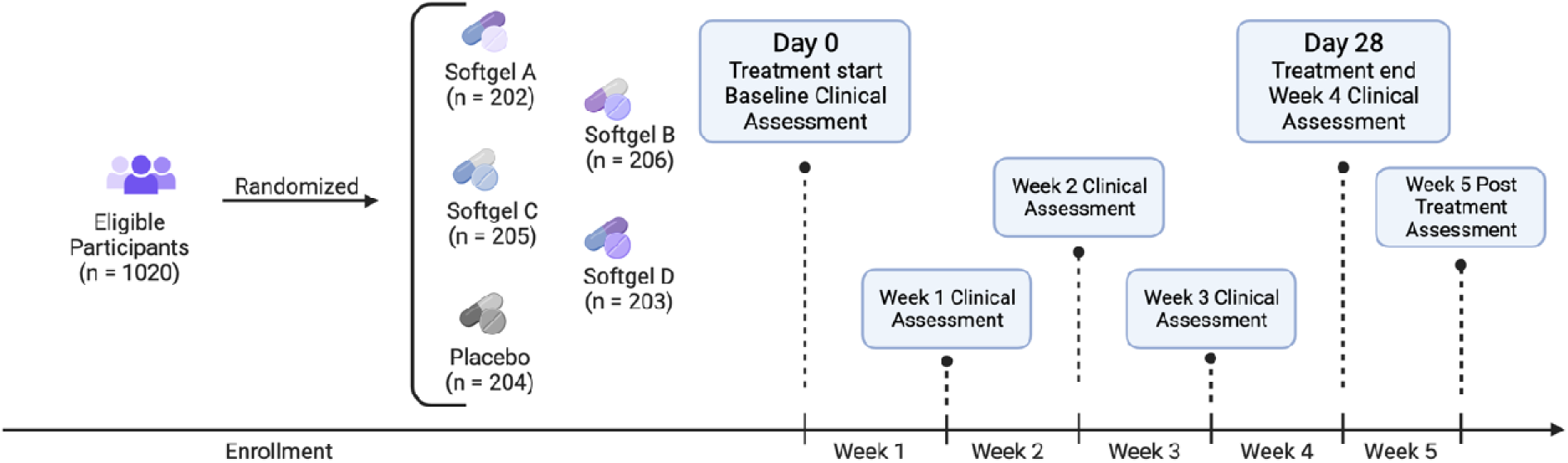
Study flow diagram. Eligible participants were enrolled in the study and randomized into one of five groups. We collected baseline clinical measures before participants started using their study product. Participants used study product for 4 weeks total. Clinical and other measures were collected at the end each week as well as 1 week post study product use.

### Randomization

Participants were randomly assigned to one of the five study product arms, with an equal chance of being assigned to each group (1:1:1:1:1 ratio). Prior to randomization, participants were stratified by their assigned sex at birth (male, female) then randomized to one of the study arms using the randomizer with evenly presented elements in the Qualtrics® XM platform.

### Outcomes

The primary focus of this study was to assess the change in the PROMIS™ Sleep Disturbance 8A scale as the primary outcome (power calculation provided below). Additionally, the study evaluated the odds of achieving a minimal clinically important difference (MCID), defined as a reduction equal to or greater than half of the standard deviation of the baseline score. The MCID standard deviation criterion was calculated separately for each study arm.

As for secondary outcomes, the study examined changes in anxiety, stress, pain, and overall well-being. The secondary outcome assessment also included monitoring the number, type, severity, causality, and outcome of side effects, adverse events, and unanticipated problems for each study arm and for the overall study.

The change in sleep quality between the active and placebo product groups was evaluated using the PROMIS Sleep Disturbance 8A scale. Secondary outcomes were assessed using the following instruments: WHO 5 for overall well-being, PROMIS™ Anxiety 4A for anxiety levels, PEG for pain intensity and interference, and PROMIS Stress 4A for stress levels.

### Safety

The assessment of spontaneously reported side effects in this study involved examining their frequency and severity. The severity of side effects was determined based on the utilization of medical services reported in response to the side effects. A grading schema was employed, following the Common Terminology Criteria for Adverse Events (CTCAE; v5.0 USDHHS): *mild*: no intervention (medication or medical advice) needed; *moderate*: a medication was taken due to the side effect or a participant sought medical advice from their HCP, urgent clinic or ED; *severe*: the side effect was medically significant but not life-threatening and/or the participant was admitted to the hospital for care and attention; *life threatening*: immediate medical intervention required and the participant was hospitalized, placed in the intensive care unit due the side effect, and/or suffered long-lasting negative effects as a result of the side effect.

### Covariates

Prior to conducting the analysis, three demographic variables (race, education, and ethnicity) were collapsed for simplicity. Race was recoded into categories: white, non-white (including participants identifying as Black, Multi-racial, Asian, some other race, American Indian or Alaska Native, or Native Hawaiian or Pacific Islander), and Prefer not to say. Education was recoded as either having a college degree (including participants with a bachelor’s or associate degree, and masters or professional degree) or no college degree (including participants with less than high school, trade/technical/vocational degree, high school diploma without college, and some college without a degree). Ethnicity was recoded into categories: Hispanic, Non-Hispanic, and Prefer not to say. Baseline demographic variables, including age, recoded race, recoded ethnicity, recoded education level, sex assigned at birth (male, female), and body mass index (BMI; calculated from self-reported height and weight) were adjusted for in the analysis.

### Power analysis

To ensure adequate statistical power, a power analysis was performed using a general covariance structure and standard deviation to detect a meaningful difference in the change in our primary outcome in each study group relative to the placebo control. It was determined that a sample size of 198 participants in each study group would provide 85% power to detect a medium effect (Cohen’s d = 0.5) between each group relative to placebo, with a two-sided p-value of 0.05 (corrected for multiple comparisons using Bonferroni). To account for conservative anticipated attrition levels (45%), recruiting up to 300 participants per study arm was planned to maintain an adequate sample size.

### Statistical analysis

A linear, mixed-effects regression model was used to assess the differences in the change in the variables of interest between each active product arm versus placebo. The parameter "na.action = na.omit" was set for each model, meaning that participants were excluded only from those models for which they did not have available data. All models were fit using an unstructured covariance matrix with a random-intercept at the participant level, and a random-slope at the study week level. The models tested the difference in the interaction between product arm and study week for active arm versus placebo, controlling for sex, age, race, ethnicity, and BMI. Post hoc Bonferroni-adjusted pairwise comparisons were used to assess the differences in the interaction between TruCBN™ formulations products and 4mg melatonin. Post hoc Bonferroni-adjusted pairwise comparisons were also used to assess the differences in the odds of achieving a MCID for sleep between each active product arm placebo, controlling for sex, age, race, ethnicity, education, and BMI.

### Software

The Python programming language, version 3.95 (packages: pandas, version 1.4.3, and numpy, version 1.20.2) were used for data processing. R, version 4.2.3 (packages: nlme, version 3.1-162, marginal effects, version 0.11.1, and tidyverse, version 2.0.0) was used to conduct the statistical analyses, and package tableone version 0.13.2 was used to create table one.

## Results

### Participants

The study included participants with a sex distribution of 54% female and 46% male, while 80% identified their race as white. After stratification, the participant numbers in each group were as follows: Softgel A (202), Softgel B (206), Softgel C (205), Softgel D (203), and Placebo (204). There were no significant differences observed between the groups in terms of demographic or outcome variables at baseline (Table 2).

**Table 2.** Participant sample summary at baseline.

| | Softgel A | Placebo | Softgel C | Softgel B | Softgel D | t / $\chi^2$<br>p-value |
| --- | --- | --- | --- | --- | --- | --- |
| Variable | Mean (SD)/N (%) |  |  |  |  |  |
| **N** | 202 | 204 | 205 | 206 | 203 |  |
| **Age** | 43.25 (13.61) | 42.68 (12.00) | 43.97 (12.70) | 43.23 (12.93) | 43.13 (12.59) | 0.899 |
| **Race** |  |  |  |  |  | 0.387 |
| White | 174 (86.1) | 160 (78.4) | 167 (81.5) | 165 (80.1) | 153 (75.4) |  |
| **Non-White** | 26 (12.9) | 42 (20.6) | 36 (17.6) | 38 (18.4) | 47 (23.2) |  |
| **Prefer not to say** | 2 (1.0) | 2 (1.0) | 2 (1.0) | 3 (1.5) | 3 (1.5) |  |
| **Education: No College Degree** | 102 (50.5) | 88 (43.1) | 91 (44.4) | 111 (53.9) | 89 (43.8) | 0.112 |
| Sex At Birth: Male | 93 (46.0) | 93 (45.6) | 95 (46.3) | 94 (45.6) | 93 (45.8) | 1 |
| Hispanic, LatinX, or Spanish origin |  |  |  |  |  | 0.497 |
| Non-Hispanic | 183 (90.6) | 186 (91.2) | 182 (88.8) | 185 (89.8) | 183 (90.1) |  |
| **Hispanic** | 19 (9.4) | 18 (8.8) | 22 (10.7) | 18 (8.7) | 17 (8.4) |  |
| **Prefer not to say** | 0 (0.0) | 0 (0.0) | 1 (0.5) | 3 (1.5) | 3 (1.5) |  |
| **BMI** | 31.06 (8.72) | 31.26 (9.65) | 30.22 (8.25) | 29.93 (7.04) | 30.10 (7.26) | 0.357 |
| PROMIS Sleep Disturbance 8A | 29.37 (6.16) | 29.27 (6.08) | 30.03 (5.68) | 29.87 (6.52) | 30.52 (6.46) | 0.238 |
| PROMIS Anxiety 4A | 11.11 (3.38) | 10.82 (3.46) | 11.07 (3.44) | 10.87 (3.47) | 11.17 (3.36) | 0.802 |
| PROMIS Stress 4A | 13.63 (3.62) | 12.87 (3.83) | 13.50 (3.46) | 13.32 (3.81) | 13.75 (3.49) | 0.121 |
| Pain, Enjoyment, General Activity Scale | 5.80 (2.35) | 5.51 (2.38) | 5.37 (2.27) | 5.59 (2.31) | 5.17 (2.31) | 0.383 |

### Sleep

The interaction between Study Week and Softgel A, (melatonin) showed a significant negative association with change in sleep disturbance (β = -0.564, p = 0.029) (Figure 2, Table 3). This indicates that the effect of Study Week on sleep disturbance differed between the treatment groups, with participants in Softgel A experiencing a greater reduction in sleep disturbance over time compared to the placebo group. The same pattern was observed for the other treatment groups: Softgel B (β = -0.544, p = 0.030), Softgel C (β = -0.603, p = 0.018), and Softgel D (β = - 0.566, p = 0.023). Next, we computed post hoc analysis to compare the difference between active arms containing TruCBN™ and melatonin (Softgel A). Contrast analyses underwent Bonferroni correction and revealed no statistically significant differences in effects between Softgel A and Softgel B (t = 0.32, p > 0.05), Softgel A and Softgel C (t = -0.09, p > 0.05), or Softgel A and Softgel D (t = 0.06, p > 0.05). Finally, we did not observe any significant differences in the likelihood of achieving a minimum clinically important difference (MCID) between Softgel A (estimate = 1.43, 95% CI[0.7, 2.16], p = 0.434), Softgel B (estimate = 1.33, 95% CI[0.65, 2.01], p = 0.729), Softgel C (estimate = 1.46, 95% CI[0.72, 2.2], p = 0.348), or Softgel D (estimate = 1.39, 95% CI[0.7, 2.08, p = 0.498]) and placebo (42.2 %). MCID is defined as a change of one half the standard deviation of the baseline.

**Figure 2.**
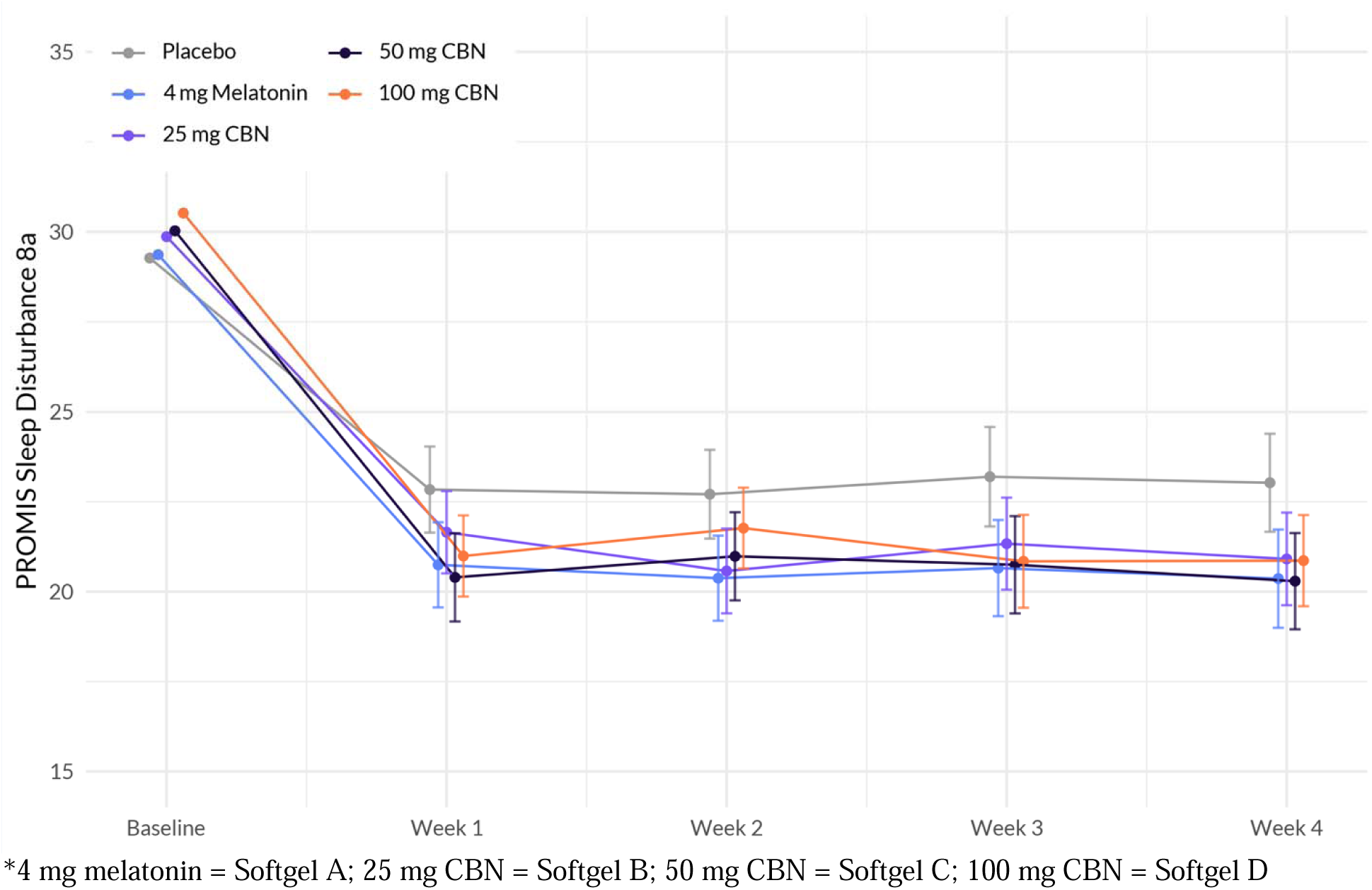
Evolution of PROMIS sleep disturbance 8a between the five arms during the study period. The plot illustrates the interaction between treatment and week on the sleep disturbance scale based on a linear mixed-effects model. The *x*-axis represents the weeks of the study, while the *y*-axis represents the outcome scale. The lines represent the trajectories of sleep disturbance for each treatment arm over time. The plot highlights the nature of treatment effects on sleep quality, as captured by the linear mixed-effects model, allowing for the incorporation of random effects and accounting for within-subject correlations.

**Table 3.** Significant Factors Associated with Sleep Disturbance: Results from a Linear Mixed Effects Regression Model. The model was used to assess the differences in the change in the variables of interest between each active product arm versus placebo. The table presents the beta coefficients (β), standard errors (Std.Error), degrees of freedom (DF), t-values, and p-values for each variable. Higher values indicate a stronger positive association with sleep disturbance, while lower values indicate a stronger negative association.

|  | Value | Std.Error | DF | t-value | p-value |
| --- | --- | --- | --- | --- | --- |
| **(Intercept)** | **25.082** | **1.036** | **1857** | **24.213** | **<0.001** |
| **Education: No College Degree [Ref: College Degree]** | **1.499** | **0.364** | **1006** | **4.121** | **<0.001** |
| **Sex at Birth: Male [Ref: Female]** | -0.666 | 0.361 | 1006 | -1.847 | 0.065 |
| Age | -0.007 | 0.014 | 1006 | -0.468 | 0.640 |
| Race: Non-white [Ref: white] | -0.573 | 0.472 | 1006 | -1.213 | 0.226 |
| Race: Prefer Not To Say [Ref: white] | 0.160 | 1.634 | 1006 | 0.098 | 0.922 |
| **BMI** | **0.080** | **0.022** | **1006** | **3.679** | **<0.001** |
| **Hispanic, LatinX, or Spanish origin: Yes [Ref: No]** | -0.794 | 0.640 | 1006 | -1.242 | 0.215 |
| Hispanic, LatinX, or Spanish origin: Prefer Not to Say [Ref: No] | -4.009 | 2.337 | 1006 | -1.716 | 0.087 |
| **StudyWeek** | **-1.123** | **0.182** | **1857** | **-6.167** | **<0.001** |
| **Softgel A [Ref: Placebo]** | -0.583 | 0.605 | 1006 | -0.964 | 0.335 |
| Softgel C [Ref: Placebo] | 0.212 | 0.604 | 1006 | 0.351 | 0.726 |
| Softgel B [Ref: Placebo] | 0.113 | 0.601 | 1006 | 0.188 | 0.851 |
| Softgel D [Ref: Placebo] | 0.547 | 0.598 | 1006 | 0.914 | 0.361 |
| **Study Week: Softgel A** | **-0.564** | **0.258** | **1857** | **-2.188** | **0.029** |
| **Study Week: Softgel C** | **-0.603** | **0.254** | **1857** | **-2.371** | **0.018** |
| **Study Week: Softgel B** | **-0.544** | **0.250** | **1857** | **-2.176** | **0.030** |
| **Study Week: Softgel D** | **-0.566** | **0.249** | **1857** | **-2.275** | **0.023** |

### Anxiety

The analysis revealed several associations with anxiety (Figure 3, Table 4). Education demonstrated a significant positive association with change in anxiety (β = 1.141, p < 0.001), indicating that individuals without a college degree reported higher levels of anxiety. BMI also exhibited a significant positive association with anxiety (β = 0.032, p = 0.007), suggesting that higher BMI was associated with higher levels of anxiety. However, the interactions between Study Week and Softgel A (Arm 1) (β = -0.095, p = 0.490), Softgel B (β = -0.028, p = 0.838), Softgel C (β = 0.130, p = 0.346), and Softgel D (β = -0.232, p = 0.086) did not reach statistical significance at the 0.05 level. Therefore, the effect of Study Week on anxiety did not significantly differ between the treatment groups compared to placebo (Figure 3, Table 4).

**Figure 3.**
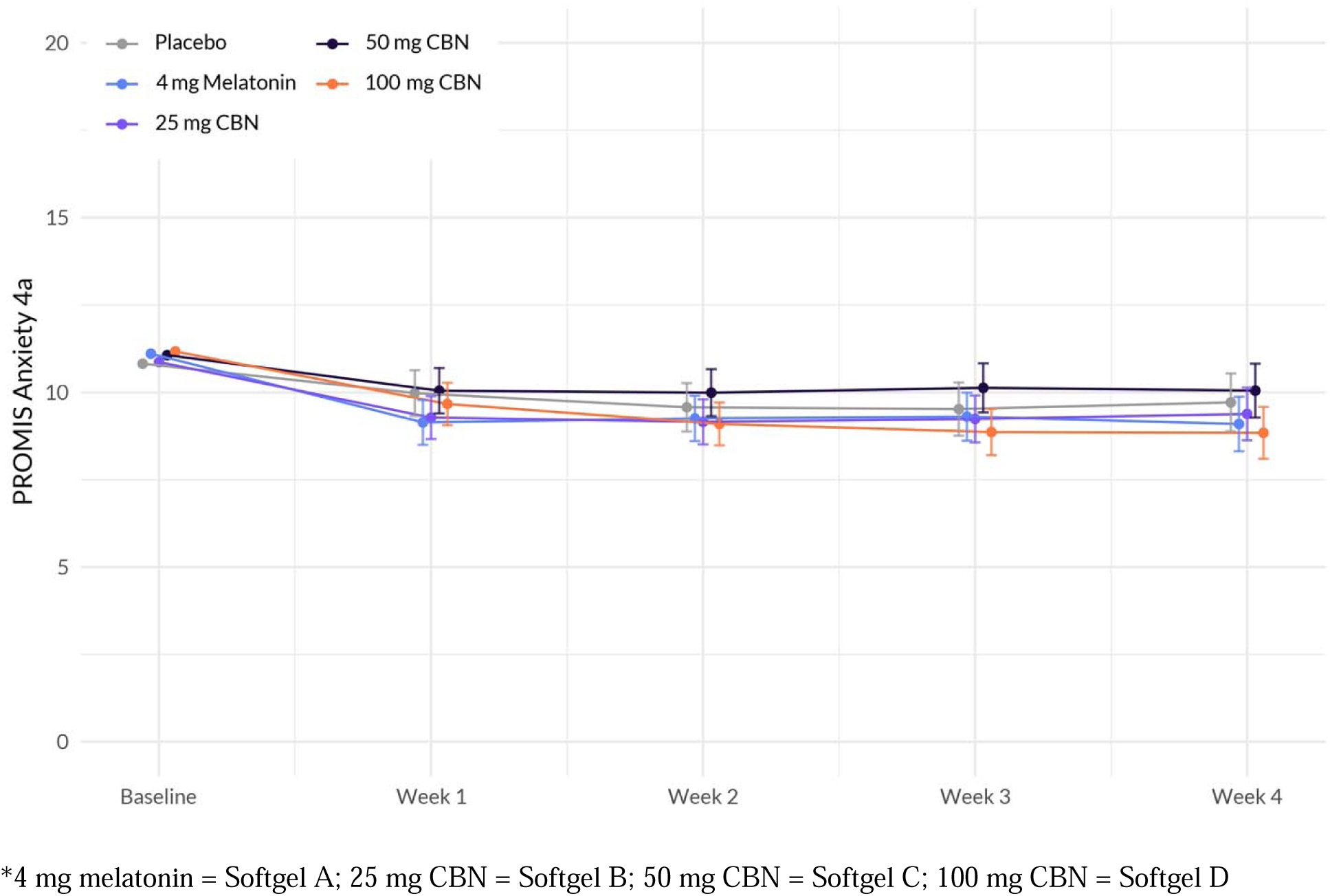
Evolution of PROMIS Anxiety 4a scores between the five arms during the study period. The plot illustrates the interaction between treatment and week on the anxiety scale, based on a linear mixed-effects model. The *x*-axis represents the weeks of the study, while the *y*-axis represents the outcome scale. The lines represent the trajectories of anxiety for each treatment arm over time. The plot highlights the nature of treatment effects on anxiety as captured by the linear mixed-effects model, allowing for the incorporation of random effects and accounting for within-subject correlations.

**Table 4.** Significant Factors Associated with Anxiety: Results from a Linear Mixed Effects Regression Model. The model was used to assess the differences in the change in the variables of interest between each active product arm versus placebo. The table presents the beta coefficients (β), standard errors (Std.Error), degrees of freedom (DF), t-values, and p-values for each variable. Higher values indicate a stronger positive association with anxiety, while lower values indicate a stronger negative association.

|  | Value | Std.Error | DF | t-value | p-value |
| --- | --- | --- | --- | --- | --- |
| **(Intercept)** | **11.407** | **0.556** | **1190** | **20.502** | **<0.001** |
| **Education: No College Degree [Ref: College Degree]** | **1.141** | **0.197** | **1006** | **5.793** | **<0.001** |
| **Sex at Birth: Male [Ref: Female]** | -0.277 | 0.196 | 1006 | -1.414 | 0.158 |
| Age | **-0.051** | **0.008** | **1006** | **-6.561** | **<0.001** |
| **Race: Non-white [Ref: white]** | 0.088 | 0.255 | 1006 | 0.345 | 0.730 |
| Race: Prefer Not To Say [Ref: white] | 0.977 | 0.898 | 1006 | 1.087 | 0.277 |
| **BMI** | **0.032** | **0.012** | **1006** | **2.697** | **0.007** |
| Hispanic, LatinX, or Spanish origin: Yes [Ref: No] | -0.116 | 0.346 | 1006 | -0.336 | 0.737 |
| Hispanic, LatinX, or Spanish origin: Prefer Not to Say [Ref: No] | -1.289 | 1.239 | 1006 | -1.040 | 0.299 |
| StudyWeek | **-0.412** | **0.101** | **1190** | **-4.082** | **<0.001** |
| **Softgel A [Ref: Placebo]** | 0.101 | 0.315 | 1006 | 0.319 | 0.750 |
| Softgel C [Ref: Placebo] | 0.334 | 0.314 | 1006 | 1.066 | 0.287 |
| Softgel B [Ref: Placebo] | -0.169 | 0.314 | 1006 | -0.538 | 0.591 |
| Softgel D [Ref: Placebo] | 0.331 | 0.313 | 1006 | 1.057 | 0.291 |
| Study Week: Softgel A | -0.095 | 0.138 | 1190 | -0.691 | 0.490 |
| Study Week: Softgel C | 0.130 | 0.138 | 1190 | 0.943 | 0.346 |
| Study Week: Softgel B | -0.028 | 0.136 | 1190 | -0.205 | 0.838 |
| Study Week: Softgel D | -0.232 | 0.135 | 1190 | -1.720 | 0.086 |

### Stress

The analysis revealed several associations with change in stress (Figure 4, Table 5). While there was no significant interaction between Study Week and Softgel A (Arm 1) (β = -0.189, p = 0.162), Softgel B (β = -0.195, p = 0.132), and Softgel C (β = -0.228, p = 0.085), a significant negative interaction was observed between Study Week and Softgel D (β = -0.323, p = 0.011). This indicates that the effect of Study Week on stress levels differed between the treatment groups, with participants in Softgel D showing a larger decrease in stress over time compared to the placebo group.

**Figure 4.**
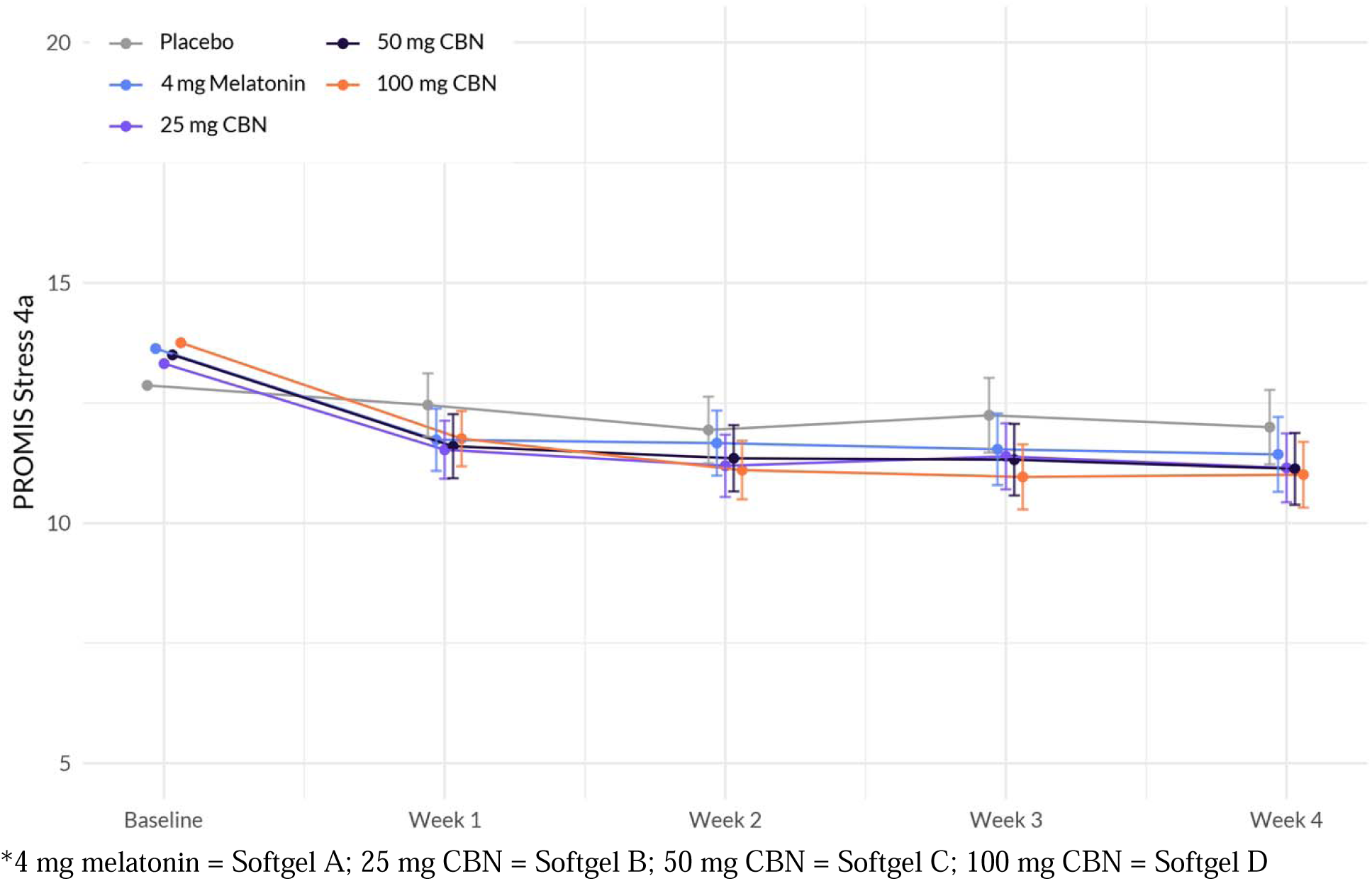
Evolution of PROMIS Stress 4a between the five arms during the study period. The plot illustrates the interaction between treatment and week on the stress scale, based on a linear mixed-effects model. The *x*-axis represents the weeks of the study, while the *y*-axis represents the outcome scale. The lines represent the trajectories of stress for each treatment arm over time. The plot highlights the nature of treatment effects on stress as captured by the linear mixedeffects model, allowing for the incorporation of random effects and accounting for within-subject correlations.

**Table 5.** Significant Factors Associated with Stress: Results from a Linear Mixed Effects Regression Model. The model was used to assess the differences in the change in the variables of interest between each active product arm versus placebo. The table presents the beta coefficients (β), standard errors (Std.Error), degrees of freedom (DF), t-values, and p-values for each variable. Higher values indicate a stronger positive association with stress, while lower values indicate a stronger negative association.

|  | Value | Std.Error | DF | t-value | p-value |
| --- | --- | --- | --- | --- | --- |
| **(Intercept)** | **14.056** | **0.591** | **1256** | **23.797** | **<0.001** |
| **Education: No College Degree [Ref: College Degree]** | **0.974** | **0.209** | **1006** | **4.669** | **<0.001** |
| **Sex at Birth: Male [Ref: Female]** | **-0.715** | **0.207** | **1006** | **-3.448** | **0.001** |
| **Age** | **-0.069** | **0.008** | **1006** | **-8.416** | **<0.001** |
| **Race: Non-white [Ref: white]** | -0.195 | 0.270 | 1006 | -0.723 | 0.470 |
| Race: Prefer Not To Say [Ref: white] | 1.150 | 0.937 | 1006 | 1.228 | 0.220 |
| **BMI** | **0.050** | **0.013** | **1006** | **3.984** | **<0.001** |
| **Hispanic, LatinX, or Spanish origin: Yes [Ref: No]** | -0.433 | 0.363 | 1006 | -1.193 | 0.233 |
| **Hispanic, LatinX, or Spanish origin: Prefer Not to Say [Ref: No]** | -1.906 | 1.285 | 1006 | -1.483 | 0.138 |
| **StudyWeek** | **-0.353** | **0.095** | **1256** | **-3.699** | **<0.001** |
| **Softgel A [Ref: Placebo]** | 0.612 | 0.333 | 1006 | 1.840 | 0.066 |
| Softgel C [Ref: Placebo] | 0.637 | 0.331 | 1006 | 1.926 | 0.054 |
| Softgel B [Ref: Placebo] | 0.257 | 0.330 | 1006 | 0.777 | 0.437 |
| **Softgel D [Ref: Placebo]** | **0.776** | **0.330** | **1006** | **2.353** | **0.019** |
| Study Week: Softgel A | -0.189 | 0.135 | 1256 | -1.400 | 0.162 |
| Study Week: Softgel C | -0.228 | 0.132 | 1256 | -1.723 | 0.085 |
| Study Week: Softgel B | -0.195 | 0.129 | 1256 | -1.508 | 0.132 |
| **Study Week: Softgel D** | **-0.323** | **0.127** | **1256** | **-2.537** | **0.011** |

### Pain

We did not detect any significant interactions between Study Week and Softgel A (Arm 1) (β = - 0.081, p = 0.401), Softgel B (β = -0.125, p = 0.1680), Softgel C (β = 0.057, p = 0.5491), and Softgel D (β = -0.119, p = 0.1888) at the 0.05 level (Figure 5, Table 6). Therefore, the effect of Study Week on pain did not significantly differ between the treatment groups compared to placebo. Several demographic variables were significantly associated with change in pain.

**Figure 5.**
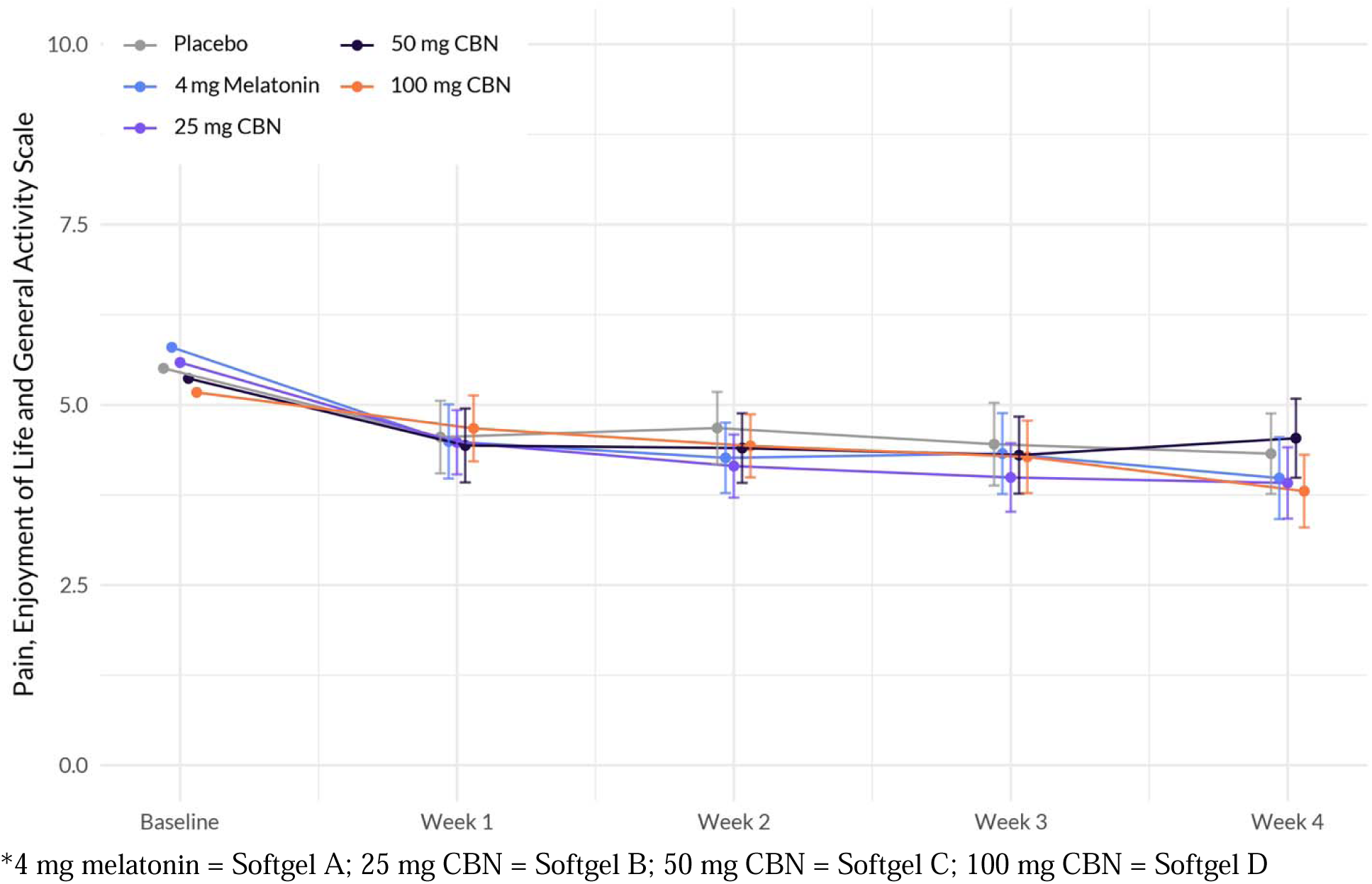
Evolution of PEG scores between the five arms during the study period. The plot illustrates the interaction between treatment and week on PEG scores, based on a linear mixedeffects model. The *x*-axis represents the weeks of the study, while the *y*-axis represents the outcome scale. The lines represent the trajectories of PEG scores for each treatment arm over time. The plot highlights the nature of treatment effects on PEG scores as captured by the linear mixed-effects model, allowing for the incorporation of random effects and accounting for within-subject correlations.

**Table 6.** Significant Factors Associated with Pain: Results from a Linear Mixed Effects Regression Model. The model was used to assess the differences in the change in the variables of interest between each active product arm versus placebo. The table presents the beta coefficients (β), standard errors (Std.Error), degrees of freedom (DF), t-values, and p-values for each variable. Higher values indicate a stronger positive association with pain, while lower values indicate a stronger negative association.

|  | Value | Std.Error | DF | t-value | p-value |
| --- | --- | --- | --- | --- | --- |
| **(Intercept)** | **2.727** | **0.561** | **952** | **4.8643** | **<0.001** |
| **Education: No College Degree [Ref: College Degree]** | **0.951** | **0.191** | **501** | **4.9786** | **<0.001** |
| **Sex at Birth: Male [Ref: Female]** | **-0.425** | **0.195** | **501** | **-2.1792** | **0.0298** |
| **Age** | **0.023** | **0.008** | **501** | **3.0401** | **0.0025** |
| Race: Non-white [Ref: white] | -0.282 | 0.265 | 501 | -1.0666 | 0.2867 |
| Race: Prefer Not To Say [Ref: white] | 0.853 | 0.883 | 501 | 0.9660 | 0.3345 |
| **BMI** | **0.040** | **0.011** | **501** | **3.7886** | **<0.001** |
| **Hispanic, LatinX, or Spanish origin: Yes [Ref: No]** | **-0.195** | **0.371** | **501** | **-0.5264** | **0.5989** |
| Hispanic, LatinX, or Spanish origin: Prefer Not to Say [Ref: No] | -0.156 | 1.088 | 501 | -0.1431 | 0.8862 |
| **StudyWeek** | **-0.216** | **0.068** | **952** | **-3.1741** | **0.0016** |
| Softgel A [Ref: Placebo] | 0.151 | 0.306 | 501 | 0.4953 | 0.6206 |
| Softgel C [Ref: Placebo] | -0.193 | 0.305 | 501 | -0.6330 | 0.5270 |
| Softgel B [Ref: Placebo] | -0.024 | 0.300 | 501 | -0.0806 | 0.9358 |
| Softgel D [Ref: Placebo] | -0.156 | 0.299 | 501 | -0.5221 | 0.6018 |
| Study Week: Softgel A | -0.081 | 0.096 | 952 | -0.8402 | 0.4010 |
| Study Week: Softgel C | 0.057 | 0.094 | 952 | 0.5994 | 0.5491 |
| Study Week: Softgel B | -0.125 | 0.090 | 952 | -1.3797 | 0.1680 |
| Study Week: Softgel D | -0.119 | 0.091 | 952 | -1.3149 | 0.1888 |

### Overall Well being

We did not detect any significant interactions between Study Week and Softgel A (Arm 1) (β = 0.302, p = 0.079), Softgel B (β = 0.280, p = 0.093), Softgel C (β = 0.194, p = 0.252), and Softgel D (β = 0.243, p = 0.143) at the 0.05 level (Figure 6, Table 7). Therefore, the effect of Study Week on change in overall wellbeing did not significantly differ between the treatment groups compared to placebo.

**Figure 6.**
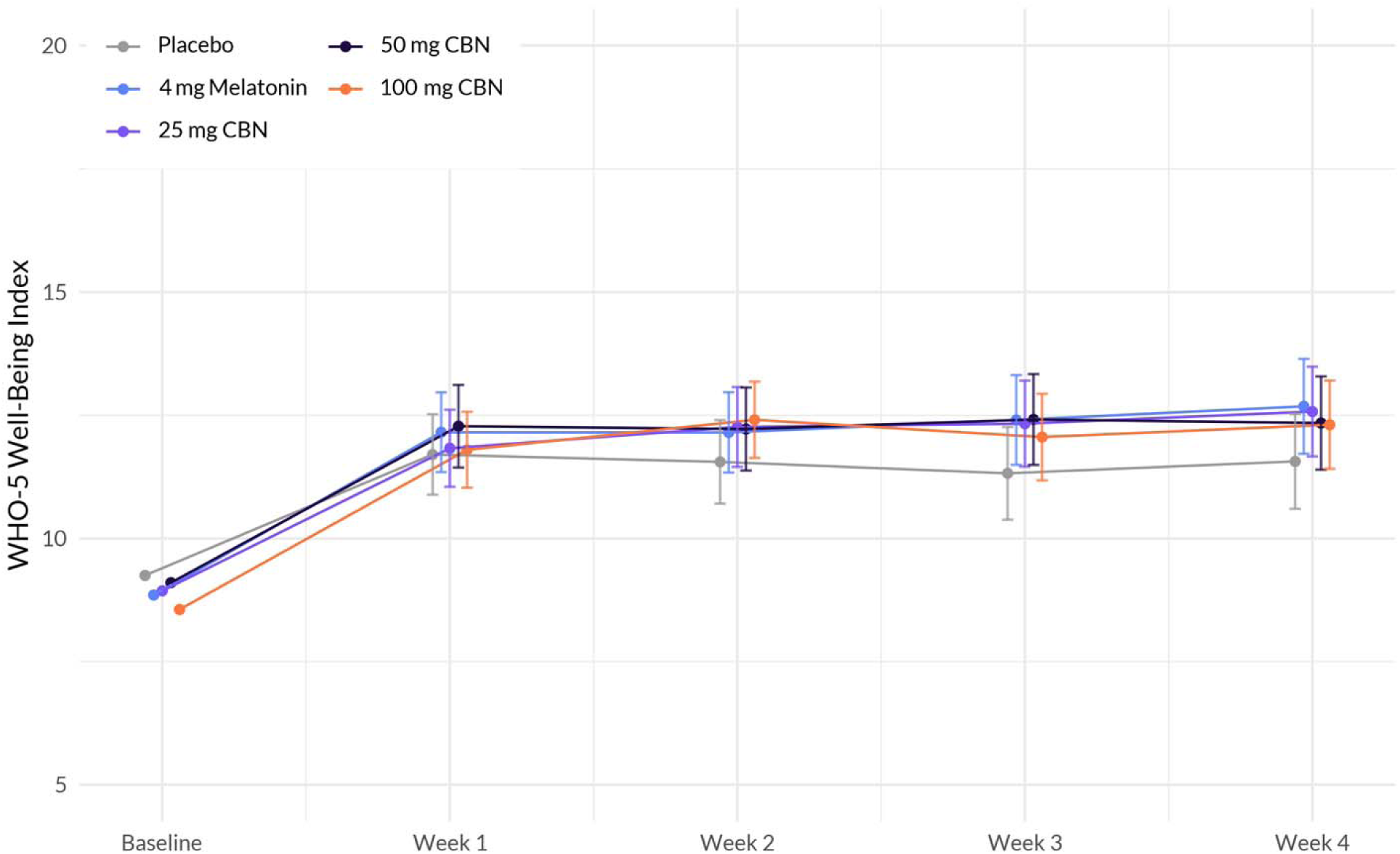
Evolution of WHO well-being scores between the five arms during the study period. The plot illustrates the interaction between treatment and week on the well-being scale, based on a linear mixed-effects model. The *x*-axis represents the weeks of the study, while the *y*-axis represents the outcome scale. The lines represent the trajectories of well-being for each treatment arm over time. The plot highlights the nature of treatment effects on well-being as captured by the linear mixed-effects model, allowing for the incorporation of random effects and accounting for within-subject correlations.

**Table 7.** Significant Factors Associated with Overall Well-Being: Results from a Linear Mixed Effects Regression Model. The model was used to assess the differences in the change in the variables of interest between each active product arm versus placebo. The table presents the beta coefficients (β), standard errors (Std.Error), degrees of freedom (DF), t-values, and p-values for each variable. Higher values indicate a stronger positive association with overall well-being, while lower values indicate a stronger negative association.

|  | Value | Std.Error | DF | t-value | p-value |
| --- | --- | --- | --- | --- | --- |
| **(Intercept)** | 10.832 | 0.765 | 1857 | 14.151 | <0.001 |
| **Education: No College Degree [Ref: College Degree]** | **-1.541** | **0.271** | **1006** | **-5.692** | **<0.001** |
| **Sex at Birth: Male [Ref: Female]** | **0.809** | **0.269** | **1006** | **3.011** | **0.003** |
| **Age** | **0.032** | **0.011** | **1006** | **2.985** | **0.003** |
| **Race: Non-white [Ref: white]** | **0.727** | **0.350** | **1006** | **2.077** | **0.038** |
| Race: Prefer Not To Say [Ref: white] | -0.221 | 1.235 | 1006 | -0.179 | 0.858 |
| **BMI** | **-0.072** | **0.016** | **1006** | **-4.428** | **<0.001** |
| Hispanic, LatinX, or Spanish origin: Yes [Ref: No] | 0.130 | 0.473 | 1006 | 0.275 | 0.783 |
| Hispanic, LatinX, or Spanish origin: Prefer Not to Say [Ref: No] | 1.941 | 1.668 | 1006 | 1.164 | 0.245 |
| **StudyWeek** | **0.457** | **0.121** | **1857** | **3.765** | **0.000** |
| Softgel A [Ref: Placebo] | -0.163 | 0.431 | 1006 | -0.378 | 0.706 |
| Softgel C [Ref: Placebo] | -0.114 | 0.429 | 1006 | -0.266 | 0.790 |
| Softgel B [Ref: Placebo] | -0.186 | 0.429 | 1006 | -0.433 | 0.665 |
| Softgel D [Ref: Placebo] | -0.678 | 0.428 | 1006 | -1.586 | 0.113 |
| Study Week: Softgel A | 0.302 | 0.172 | 1857 | 1.756 | 0.079 |
| Study Week: Softgel C | 0.194 | 0.169 | 1857 | 1.147 | 0.252 |
| Study Week: Softgel B | 0.280 | 0.167 | 1857 | 1.679 | 0.093 |
| Study Week: Softgel D | 0.243 | 0.166 | 1857 | 1.464 | 0.143 |

### Side effects

There was no significant difference in the frequency of reported side effects between the placebo group and the other groups (χ² (4) = 8.58, p = 0.073)). Participants in the study reported a range of side effects, including Grogginess/drowsiness (n=22), Difficulty falling or staying asleep (n=10), Headache (n=10), Nausea (n=5), Nightmares (n=5), Diarrhea (n=4), Upset stomach (n=3), Strange or vivid dreams (n=3), Anxious or restless feelings (n=3), Hallucinations (n=2), Racing heart (n=2), and Gas/bloating (n=2). Additionally, each of the following side effects was reported by one participant each: Chest pain, Constipation, Urinary Tract Infection, tingling in lips, Dry mouth, Weight gain, Dry or itchy eyes, Low Libido, Rash, and Brain fog. Side effects were mild; none was considered serious or required use of emergency or non-emergency healthcare services (Figure 7).

**Figure 7.**
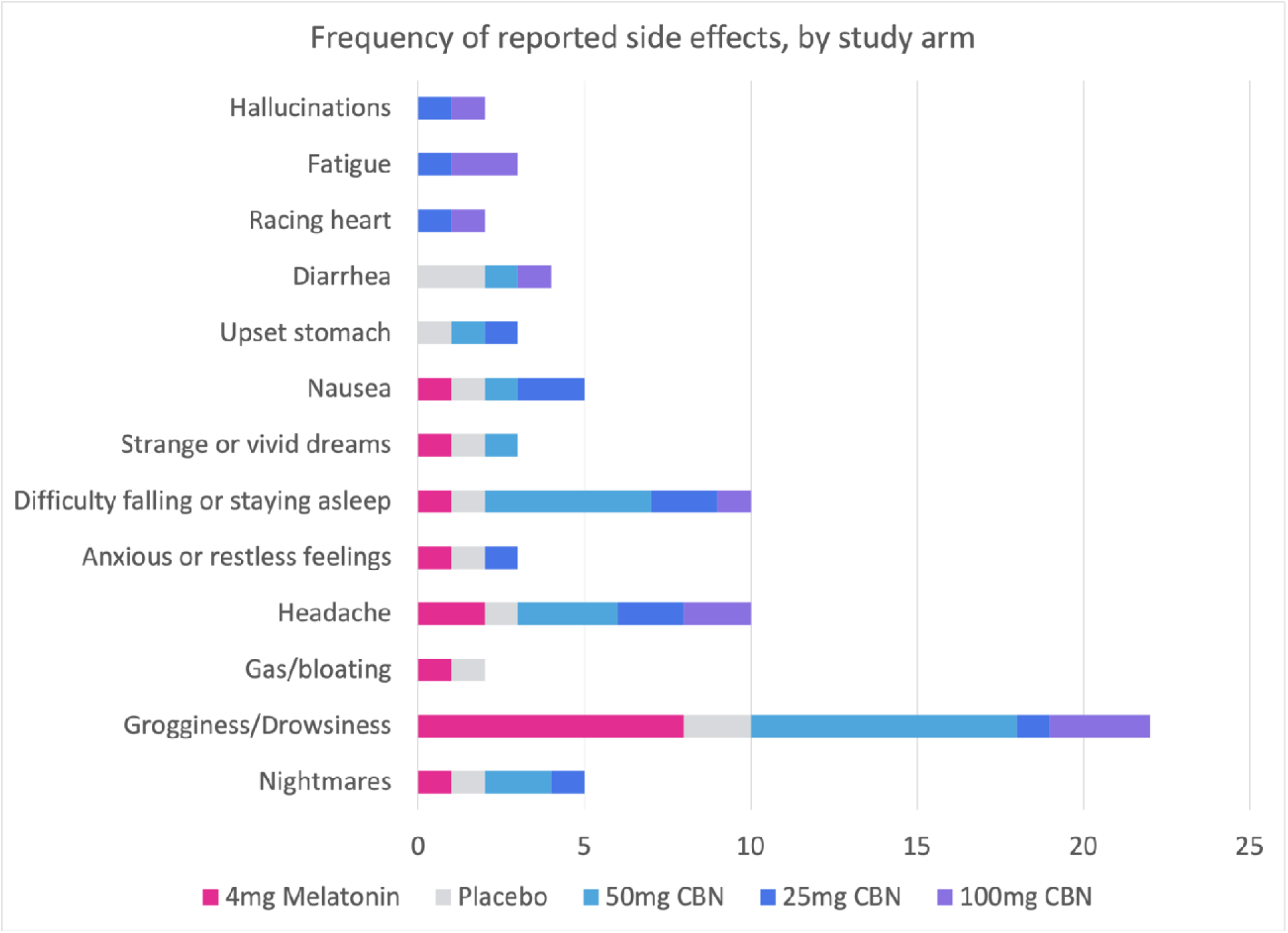
Comparison of side effects between active and placebo. This figure displays the occurrence of side effects in the active and placebo groups. All side effects were mild and nonserious, requiring no emergency or non-emergency healthcare services.

## Discussion

The primary analyses of this study revealed significant improvements in sleep outcomes as measured by the PROMIS Sleep Disturbance 8a score. The key findings were a significant difference in the rate of mean PROMIS Sleep Disturbance 8a score change between all groups relative to placebo, indicating that the 25, 50 and 100 mg serving sizes of TruCBN™ were effective in improving sleep disturbances. Additionally, the group taking 4 mg of melatonin reported significant improvements in sleep outcomes compared to placebo. All side effects were mild or moderate. There were no significant differences in the frequency of reported side effects between any dose of TruCBN™ formulations or melatonin compared to placebo.

While CBN is a popular alternative to prescription drugs to improve sleep outcomes, there are few well-designed clinical trials to assess its effectiveness, especially as a single ingredient. The findings of this study are consistent with previous findings that a combination of cannabinoids could improve sleep. For example, a randomized, controlled crossover trial administering a combination product, containing THC 20 mg/mL, CBN 2 mg/mL, CBD 1 mg/mL, and naturally occurring terpenes (extracted from the cannabis plant), in pharmaceutical grade sunflower oil, for 2 weeks, demonstrated an improvement in sleep quality in subjects with insomnia when compared to placebo.^24^ In a recent study we reported that a botanical blend containing a low concentration of THC, CBD and CBN improved sleep disturbance, anxiety, stress, and well-being in healthy individuals that reported better sleep as a primary health concern.^22^ In a similar study subjects receiving a tablet containing 10 mg THC and 5 mg CBN nightly experienced significantly improved sleep quality and slept significantly longer, with a 5% increase in sleep duration.^20^ We recently published the results of a similar sleep study on 1793 adults.^23^ Participants were randomly assigned to take 1 of 6 products, containing either 15 mg CBD or 5 mg melatonin, alone or in combination with minor cannabinoids, including CBN. Most participants (56% to 75%) across all formulations experienced a clinically important improvement in their sleep quality though not statistically better than the active control group that took 5 mg of melatonin alone. Our results are further supported by the role CBN plays on the ECS. However, to our knowledge this is the first study of this size to demonstrate the effectiveness of highly purified CBN isolate formulations containing 25, 50 and 100 mg doses of TruCBN™ for improvement in sleep outcomes.

This study was intended to assess the “real world” effectiveness of varying doses of CBN by administering them to a broad population representative of a consumer seeking such a product for sleep disturbances and the other outcomes studied. This is in contrast to traditional clinical trials, which often have restrictive eligibility criteria, rigorous monitoring and are limited to those who can access the site being utilized to conduct the trial. As a result, traditional trials often exhibit higher levels of missingness and heterogeneity and lack external validity, as the participants’ characteristics and behaviors may not accurately represent those of real-world users. It was the goal of this study to reflect the real-world effects of the study products and as a distinct and unique way to provide evidence for regulatory and clinical decision-making and additional clinical trial design.^25^

The study has a few limitations. First, approximately 26% of participants did not complete any follow-up surveys. However, the overall attrition level was still below our anticipated attrition (45%) and the study remained adequately powered to detect significant sleep changes. We were also unable to perform a sensitivity analysis including excluded participants as they did not provide any PROMIS Sleep Disturbance 8a scores. Thus, including their data would not be appropriate as their data were determined to be missing not at random. Nevertheless, there were no significant differences in baseline demographic or health characteristics between study arms in the final study sample. This indicates that balance was maintained across study arms. In addition, covariate regression analysis was performed to further account for potential sources of confounding.

## Conclusion

The results demonstrate that TruCBN™, consumed orally in a softgel with serving sizes of 25 mg, 50 mg and 100 mg, could be a safe and effective alternative therapy for the improvement of sleep. There was no significant difference in improvements in sleep quality between any of the treatment groups and 4mg melatonin, indicating that TruCBN™ offers an alternative for effective sleep support.

## Data Availability

All data produced in the present work are contained in the manuscript

## Acknowledgments

The authors would like to express our deepest gratitude to the participant volunteers in this study without whom this study would not have been possible. We acknowledge and appreciate the hard work of the Radicle Science study operations team and FloraWorks for the product formulas studied.

## Author Contributions

Conceptualization and Methodology, J. Chen and E. Pauli. Data curation and formal analysis, C. Bryant.; writing—original draft preparation, A. Kolobaric, J. Saleska and S.Hewlings.; writing—review and editing, all authors. All authors have read and agreed to the published version of the manuscript.

## References

1. Havekes, R., Park, A., Tudor, J., Luczak, V., Hansen, R., Abel, T., … & Baillie, G. (2016). Sleep Deprivation Causes Memory Deficits By Negatively Impacting Neuronal Connectivity In Hippocampal Area Ca1. Elife, (5). 10.7554/elife.13424

2. Alshabibi???? Durmer, J., Dinges, D. (2005). Neurocognitive Consequences Of Sleep Deprivation. Semin Neurol, 01(25), 117–129. 10.1055/s-2005-867080

3. Freitag, L., Ireland, J., Niesten, I. (2017). Exploring the Relationship Between Sleep Quality, Emotional Well-being And Aggression Levels In A European Sample. JACPR, 3(9), 167–177. 10.1108/jacpr-08-2016-0239

4. Son, S., Park, E., Cho, Y., Lee, S., Choi, J., Lee, Y., … & Kim, C. (2020). <p>association Between Weekend Catch-up Sleep and Metabolic Syndrome With Sleep Restriction In Korean Adults: A Cross-sectional Study Using Knhanes</p>. DMSO, (Volume 13), 1465-1471. 10.2147/dmso.s247898

5. Weil, Z. M., Norman, G. J., Karelina, K., Morris, J. S., Barker, J. M., Su, A., … & DeVries, A. C. (2009). Sleep Deprivation Attenuates Inflammatory Responses and Ischemic Cell Death. Experimental Neurology, 1(218), 129–136. 10.1016/j.expneurol.2009.04.018

6. Scott AJ, Webb TL, Martyn-St James M, Rowse G, Weich S. Improving sleep quality leads to better mental health: A meta-analysis of randomised controlled trials. Sleep Med Rev. 2021;60:101556. doi:10.1016/j.smrv.2021.101556

7. Institute of Medicine Committee on Sleep, M.; Research. The National Academies Collection: Reports funded by National Institutes of Health. In Sleep Disorders and Sleep Deprivation: An Unmet Public Health Problem, Colten, H.R., Altevogt, B.M., Eds.; National Academies Press (US) Copyright © 2006, National Academy of Sciences.: Washington (DC), 2006.

8. Adjaye-Gbewonyo D, Ng AE, Black LI. Sleep difficulties in adults: United States, 2020. NCHS Data Brief, no 436. 2022; 436:1–8. Free full text article: https://stacks.cdc.gov/view/cdc/117490

9. Buysse, D. J. (2013). Insomnia. JAMA, 7(309), 706. 10.1001/jama.2013.193

10. Sateia, M. J., Buysse, D. J., Krystal, A. D., Neubauer, D., Heald, J. L. (2017). Clinical Practice Guideline For the Pharmacologic Treatment Of Chronic Insomnia In Adults: An American Academy Of Sleep Medicine Clinical Practice Guideline. Journal of Clinical Sleep Medicine, 02(13), 307–349. 10.5664/jcsm.6470

11. Atkin T, Comai S, Gobbi G. Drugs for Insomnia beyond Benzodiazepines: Pharmacology, Clinical Applications, and Discovery. Pharmacol Rev. 2018 Apr;70(2):197–245. doi: 10.1124/pr.117.014381. PMID: 29487083.

12. Fatemeh G, Sajjad M, Niloufar R, Neda S, Leila S, Khadijeh M. Effect of melatonin supplementation on sleep quality: a systematic review and meta-analysis of randomized controlled trials. J Neurol. 2022;269(1):205–216. doi:10.1007/s00415-020-10381-w

13. Chan V, Lo K. Efficacy of dietary supplements on improving sleep quality: a systematic review and meta-analysis. Postgrad Med J. 2022;98(1158):285–293. doi:10.1136/postgradmedj-2020-139319

14. Li T, Jiang S, Han M, et al. Exogenous melatonin as a treatment for secondary sleep disorders: A systematic review and meta-analysis. Front Neuroendocrinol. 2019;52:22–28. doi:10.1016/j.yfrne.2018.06.004

15. Choi S, Huang BC, Gamaldo CE. Therapeutic Uses of Cannabis on Sleep Disorders and Related Conditions. J Clin Neurophysiol. 2020 Jan;37(1):39–49. doi: 10.1097/WNP.0000000000000617. Erratum in: J Clin Neurophysiol. 2020 Sep;37(5):466-467. PMID: 31895189.

16. Babson, K.A.; Sottile, J.; Morabito, D. Cannabis, Cannabinoids, and Sleep: a Review of the Literature. Current psychiatry reports 2017, 19, 23, doi:10.1007/s11920-017-0775-9.

17. Maldonado R, Cabañero D, Martín-García E. The endocannabinoid system in modulating fear, anxiety, and stress. Dialogues Clin Neurosci. 2020 Sep;22(3):229–239. doi: 10.31887/DCNS.2020.22.3/rmaldonado. PMID: 33162766; PMCID: PMC7605023.

18. Suraev, A.S.; Marshall, N.S.; Vandrey, R.; McCartney, D.; Benson, M.J.; McGregor, I.S.; Grunstein, R.R.; Hoyos, C.M. Cannabinoid therapies in the management of sleep disorders: A systematic review of preclinical and clinical studies. Sleep medicine reviews 2020, 53, 101339, doi:10.1016/j.smrv.2020.101339.

19. Tham, M.; Yilmaz, O.; Alaverdashvili, M.; Kelly, M.E.M.; Denovan-Wright, E.M.; Laprairie, R.B. Allosteric and orthosteric pharmacology of cannabidiol and cannabidiol-dimethylheptyl at the type 1 and type 2 cannabinoid receptors. British journal of pharmacology 2019, 176, 1455–1469, doi:10.1111/bph.14440.

20. Gannon, W.; Bronfein, W.; Jackson, D.; Holshouser, K.; Artman, B.E.; Schestepol, M.; Treacy, D.J.; Rudnic, E.M. Novel 569 Formulation of THC and CBN in a Repeat-Action Tablet Improves Objective and Subjective Measurements of Sleep. Am J 570 Endocannabinoid Medicine 2021, 3, 12–18.

21. Yoshida, H., Usami, N., Ohishi, Y., Watanabe, K., Yamamoto, I., Yoshimura, H. (1995). Synthesis and Pharmacological Effects In Mice Of Halogenated Cannabinol Derivatives.. CHEMICAL & PHARMACEUTICAL BULLETIN, 2(43), 335–337. 10.1248/cpb.43.335

22. Kolobaric A, Hewlings SJ, Bryant C, Colwell CS, R. D’Adamo C, Rosner B, Chen J, Pauli EK. A Randomized, Double-Blind, Placebo-Controlled Decentralized Trial to Assess Sleep, Health Outcomes, and Overall Well-Being in Healthy Adults Reporting Disturbed Sleep, Taking a Melatonin-Free Supplement. Nutrients. 2023; 15(17):3788. 10.3390/nu15173788

23. Saleska, J.L.; Bryant, C.; Kolobaric, A.; D’Adamo, C.R.; Colwell, C.S.; Loewy, D.; Chen, J.; Pauli, E.K. The Safety and 629 Comparative Effectiveness of Non-Psychoactive Cannabinoid Formulations for the Improvement of Sleep: A 630 Double-Blinded, Randomized Controlled Trial. Journal of the American Nutrition Association 2023, 1–11, 631 doi:10.1080/27697061.2023.2203221

24. Walsh, J.H.; Maddison, K.J.; Rankin, T.; Murray, K.; McArdle, N.; Ree, M.J.; Hillman, D.R.; Eastwood, P.R. Treating 572 insomnia symptoms with medicinal cannabis: a randomized, crossover trial of the efficacy of a cannabinoid medicine 573 compared with placebo. Sleep 2021, 44, doi:10.1093/sleep/zsab149

25. Sherman, R.E.; Anderson, S.A.; Dal Pan, G.J.; Gray, G.W.; Gross, T.; Hunter, N.L.; LaVange, L.; Marinac-Dabic, D.; Marks, 633 P.W.; Robb, M.A.; et al. Real-World Evidence - What Is It and What Can It Tell Us? The New England journal of medicine 634 2016, 375, 2293–2297, doi:10.1056/NEJMsb1609216

